# ADESSO: a rapid, adaptable and sensitive Cas13-based COVID-19 diagnostic platform

**DOI:** 10.1101/2021.06.17.21258371

**Authors:** Beatrice Casati, Joseph Peter Verdi, Alexander Hempelmann, Maximilian Kittel, Andrea Gutierrez Klaebisch, Bianca Meister, Sybille Welker, Sonal Asthana, Pavle Boskovic, Ka Hou Man, Meike Schopp, Paul Ginno, Bernhard Radlwimmer, Charles Erec Stebbins, Thomas Miethke, Fotini Nina Papavasiliou, Riccardo Pecori

## Abstract

SARS-CoV-2 is the causative agent for a pandemic that has had immense consequences for the health and economic sectors worldwide. While PCR testing and later antigen tests have proven critical for helping to stem the spread of the virus, these methods suffer from general applicability and sensitivity, respectively. Moreover, the emergence of variant strains creates the need for flexibility to correctly and efficiently diagnose the presence of substrains in the population. To address these needs we have developed the diagnostic test ADESSO (Accurate Detection of Evolving SARS-CoV-2 through SHERLOCK Optimization) which employs the Cas13 system to diagnose patients in as little as 1 hour without sophisticated equipment. Using an extensive panel of clinical samples, we demonstrate that ADESSO correctly identifies COVID-19 positive samples at a sensitivity and specificity comparable to RT-qPCR on extracted RNA and higher than antigen tests for unextracted samples. Taken together, ADESSO is a fast, sensitive and cheap method that can be applied in a point of care (POC) setting to diagnose COVID-19 and can be quickly adjusted to detect new variants.

## INTRODUCTION

Since the beginning of the coronavirus disease 2019 (COVID-19) pandemic, 228 million confirmed cases, including 4.6 million deaths, have been reported globally^1^. COVID-19 is a respiratory disease caused by severe acute respiratory syndrome coronavirus 2 (SARS-CoV-2)^2–4^. The quick diffusion of SARS-CoV-2 is primarily attributed to the relatively long duration of viral shedding by infected individuals, the viral load dynamics and the lengthy incubation period^5^ of 5-6 days^6,7^. The viral load peaks soon after the onset of symptoms^8–11^, suggesting that individuals with COVID-19 begin viral shedding a few days before symptoms appear^12^. Further, a significant proportion of infected individuals either remain entirely asymptomatic or only manifest mild symptoms^13–16^, thus facilitating the spread of the virus and leading to the current pandemic situation.

The situation has been exacerbated by the fact that SARS-CoV-2 has evolved considerably. The first variant to appear carried a D614G mutation in the spike protein^17^ which is now dominant and shared between all existing variants. The virus has since accumulated multiple additional mutations in varying combinations, resulting in more transmissible and potentially more virulent variants threatening several countries worldwide^18^.

The urgent need for a prophylactic response has accelerated the development of multiple effective vaccines and more than 5.7 billion doses have already been administered^1,19–22^. However, even in the most optimistic scenario it will take time to reap the benefits of a global vaccination campaign^23,24^, especially in the poorest countries where the vaccination rate is dramatically lower^18^. Therefore, complementary efforts to limit the spread of the virus are still essential. A recent model of viral dynamics indicates that frequent testing is essential for efficient identification and isolation of carriers and containment of the pandemic^25^, highlighting the need for an accurate and reliable test that can be easily accessed, regularly performed and whose results are returned quickly.

The worldwide gold standard diagnostic test for SARS-CoV-2 infection is the reverse transcription quantitative polymerase chain reaction (RT-qPCR)^26^. While sensitive and effective, it comes with the important limitation of requiring specific equipment, laboratory infrastructures and qualified personnel. Inadequate access to such resources significantly reduces the frequency of testing. Additionally, PCR testing facilities often need days’ worth of time to report the test outcome, resulting in a long sample-to-result turnaround time. To face these challenges, different rapid tests have been implemented, such as rapid PCR and antigen-based tests. While these advancements represent significant progress in diagnosing COVID-19, rapid PCR tests still require specific equipment^27^ and antigen-based tests have lower sensitivity and specificity^28– 30^. In fact, multiple investigations on the accuracy and reliability of antigen-based tests have pointed out that their use is beneficial only for the detection of infected individuals with high viral titers^29,31–33^. Taken as a whole, there is still a need for an alternative test that is comparable to RT-qPCR in terms of sensitivity and specificity, yet faster and independent of complex instruments.

The so-called CRISPR diagnostic (CRISPR-Dx) technologies offer promising solutions to meet all these requirements^34^. The CRISPR bacterial system is capable of recognizing and cleaving foreign genetic material. Among the CRISPR associated (Cas) proteins, Cas13 and Cas12 specifically bind RNA and DNA molecules, respectively, complementary to the target-binding CRISPR RNA (crRNA). Upon target recognition, the Cas proteins cleave a reporter in *trans*, which can then be detected via different readouts^35–37^. To achieve high sensitivity, isothermal amplification methods that do not rely on sophisticated equipment, such as loop-mediated isothermal amplification (LAMP)^38^ or recombinase polymerase amplification (RPA)^39^, have been combined with Cas-mediated nucleic acid detection^36,40,41^. CRISPR-Dx technologies were quickly adapted for the detection of SARS-CoV-2^42–49^ and two of them have received emergency use authorization from the Food and Drug Administration (FDA), with use restricted to the approved laboratories^50,51^. Despite their high potential, most of these technologies require either extracted RNA^46,52–54^ or show a lower limit of detection (LoD) when performed on unextracted samples^42,44^. Finally, while CRISPR-Dx technologies were benchmarked against RT-qPCR, the analysis of their performance on clinical samples in direct comparison with antigen tests is lacking.

Here we have optimized the Cas13a-based “SHERLOCK”^40,41^ platform to develop ADESSO (Accurate Detection of Evolving SARS-CoV-2 through SHERLOCK Optimization). ADESSO demonstrates highly sensitive detection of SARS-CoV-2 and its variants directly from patient-derived material. The entire protocol is completed in approximately one hour, does not require RNA extraction or any specific equipment, approaches the detection limit of RT-qPCR at 2.5 cp/μl of SARS-CoV-2 synthetic genome and is low-cost (less than 5€ per test). Throughout our work, we extensively evaluated the real diagnostic potential of ADESSO in direct comparison to RT-qPCR and antigen testing on samples collected with two different methods (nasopharyngeal swab (NP) and gargle of saline). To ensure our sample cohort represented a relevant portion of the population that can remain undetected, we included ambulatory patients with minimal or mild symptoms as well as asymptomatic individuals who had recently contacted COVID-19 positive patients. Our study showed that ADESSO has a sensitivity comparable to RT-qPCR when applied to purified RNA. When employed directly on unextracted samples, ADESSO outperformed the rapid antigen test, demonstrating its potential as a more sensitive and reliable POC diagnostic test.

## RESULTS

### ADESSO: an optimized and highly-sensitive SHERLOCK assay

We began developing ADESSO by determining sensitivity of the Cas13-based SHERLOCK (Specific High Sensitivity Enzymatic Reporter UnLOCKing)^40^ platform on clinical samples (**Figure S1**). To increase sensitivity and reduce duration of the assay, we evaluated alternative reagents and different reaction conditions for RNA extraction, isothermal amplification of viral RNA via RT-RPA and detection of a specific RNA sequence by Cas13 (**Figure S1C)**. We assessed Cas13 activation with a fluorometer to monitor the speed of the reaction in real-time and via a lateral flow-based visual readout as an instrument-free detection method that would be used in a POC test. The fluorescence and lateral flow readouts are based on the use of two different RNA reporters, where a fluorophore (e.g., FAM) is flanked by either a quencher (for fluorescence readouts) or a biotin (for lateral flow readouts). Upon Cas13-mediated cleavage of the reporter, the fluorophore is cut from either the quencher or the biotin. For fluorescence-based readouts, cleavage of the RNA reporter releases the fluorophore from the quencher and the fluorescent signal can then be detected by a fluorometer (**Figure S1C**). In the lateral flow scenario, the resulting signal can be read on a lateral flow strip where gold-labeled antibodies against FAM are used to visualize the reporter. In a negative sample, the RNA reporter flanked by FAM and biotin is intact and is captured by a first line of streptavidin resulting in a band called “control band”. In a positive sample, the reporter is cut, releasing the FAM-containing fragment to be captured by a second line of antibodies resulting in a “test band”. The band intensity ratio between the test band and the control band thus reflects the level of Cas13 activation (**Figure S1C**). Positive samples were empirically determined to be represented by a band intensity ratio higher than 0.2. This threshold was defined based on the band intensity ratio obtained in all the negative controls and samples used in this study (n = 282 + 400; **Figure S2**) and is in agreement with previously published findings^47^.

The RNA extraction step is a time-consuming, labour-intensive and costly step for COVID-19 diagnosis (**Figure S1C**). It has furthermore been complicated by the global shortage of RNA extraction kits throughout the pandemic^55,56^. Therefore, we optimized the SHERLOCK method parameters to allow for circumvention of this step while maintaining high sensitivity. We first minimized RNase activity during sample lysis by adding RNase Inhibitors in the lysis buffer. After a 5 min incubation at 95°C in QuickExtract DNA Extraction solution (as previously shown^42^) in the presence of RNase Inhibitors, RNaseAlert was added to the sample to evaluate nuclease activity and fluorescence was measured. Notably, addition of RNase inhibitors in the lysis buffer prior to heating was sufficient to inhibit RNAse activity almost completely (**Figure 1A**). Next, we optimized the amount of RT units and the volume of sample input in the RT-RPA step using dilutions of the synthetic SARS-CoV-2 genome. We observed the best results with 6 U/μl of RT and 2.5 μl of sample input per reaction (**Figure 1B**). Additionally, we further optimized the RT-RPA step by first comparing different RT enzymes in the presence or absence of RNase H. M-MuLV showed the best sensitivity (5-2.5 cp/μl) in comparison to ProtoScript II or SuperScript III, while the addition of RNase H led to an improvement for SuperScript III only (**Figure 1C**). Secondly, we used varying final concentrations of RPA, where 1xRPA corresponds to the standard amount of RPA described in the original SHERLOCK protocol^40,57,58^ and 5xRPA corresponds to the optimal amount according to the manufacturer’s instructions. To test this, we performed our assay with different concentrations of RPA on a positive clinical sample with Ct = 29.3 (**Supplementary File 1**), which is approximately the LoD of other Cas13-based tests^44,47^. Remarkably, while we achieved a false negative with 1xRPA, the sample resulted positive for final concentrations of RPA from 2x to 5x (**Figure S3A**). Bearing in mind the cost per single test, we decided to proceed using a 2xRPA concentration. Finally, in order to optimize the Cas13 detection step, we made a ten-fold dilution of a positive RT-RPA reaction (50 cp/μl) and we performed Cas13 detection using the original concentration of Cas13/crRNA (45/22.5 nM)^40,57,58^, in comparison to higher amounts (**Figure S3B**, upper panel). A concentration of Cas13/crRNA of 90 nM each led to a faster reaction, reaching the plateau after 15 min only, compared to 30 min for the other two concentrations (**Figure S3B**, lower panel). We also confirmed that a 10 min incubation for Cas13 detection is sufficient to yield a clearly positive outcome in the lateral flow detection assay (**Figure S3C**), which is an essential feature for a POC test. Moreover, a shorter Cas13 reaction allows us to extend the incubation time of the RT-RPA step for highly sensitive reactions^57^ without affecting the total time of the assay. Finally, we assessed the sensitivity of this optimized protocol on serial dilutions of SARS-CoV-2 synthetic genome and we observed a significant reproducible sensitivity of 2.5 cp/μl (**Figure 1D**). We named this new optimized diagnostic assay ADESSO (Accurate Detection of Evolving SARS-CoV-2 through SHERLOCK Optimization) (**Figure 1E**).

**Figure 1:**
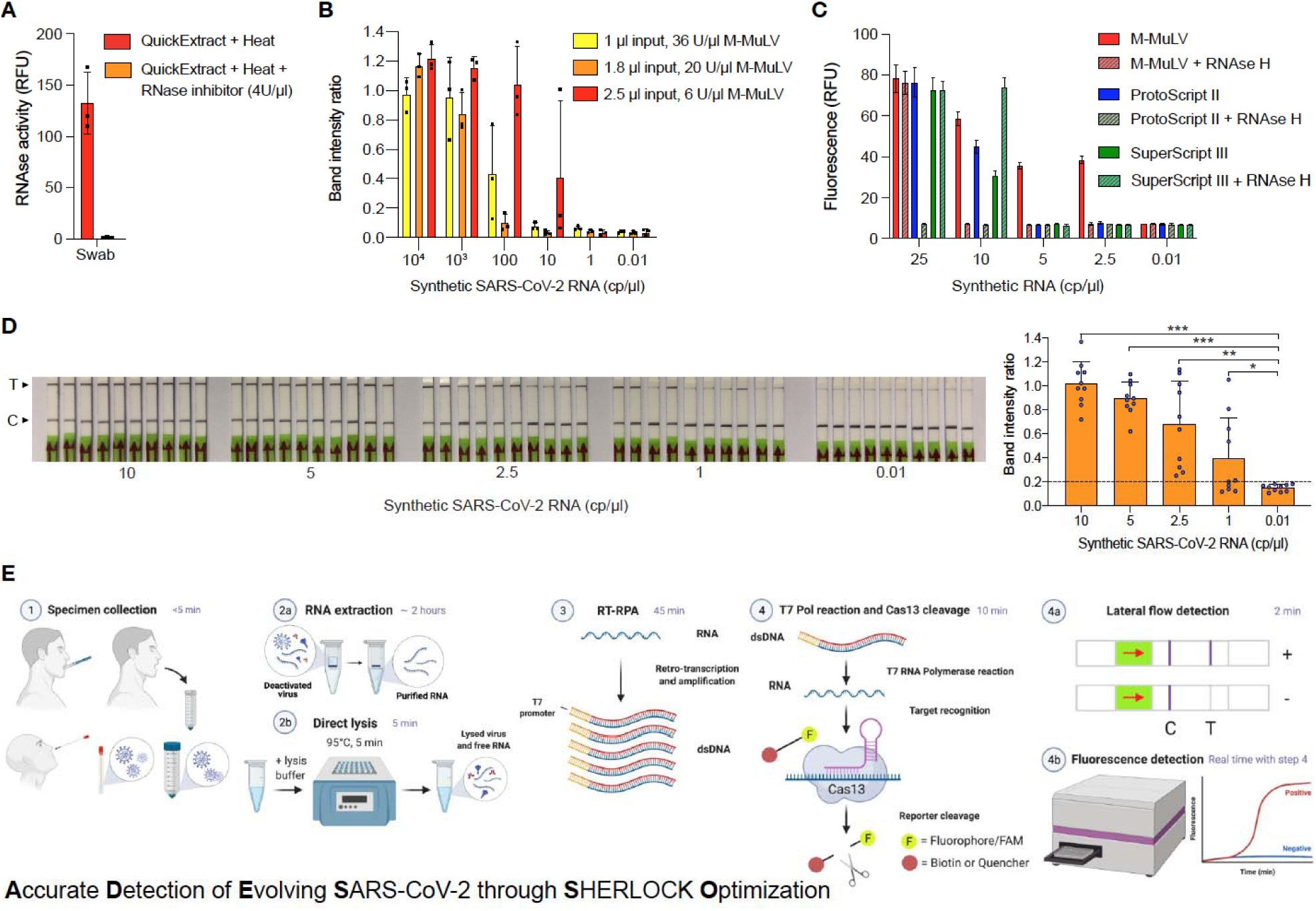
ADESSO: an optimised and highly sensitive SHERLOCK-based assay for SARS-CoV-2 detection. **A**. Measurement of RNase activity in a swab sample lysed at 95°C for 5 minutes with QuickExtract DNA Extraction Solution enriched or not with RNase inhibitor, Murine, at a final concentration of 4 U/μl. **B**. Determination of sensitivity on serial dilutions of SARS-CoV-2 synthetic genome upon optimisation of RT units and input volume in the RT-RPA reaction with lateral flow readout. **C**. Sensitivity on serial dilutions of SARS-CoV-2 genome using different reverse transcriptases in presence or absence of RNase H. **D**. Sensitivity of the improved protocol with lateral flow readout on serial dilutions of SARS-CoV-2 synthetic genome upon integration of all the above-described optimisations. Intensity ratios are shown in the bar plot on the right. **E**. Graphic of the experimental workflow of ADESSO to detect SARS-CoV-2, with or without RNA extraction, in clinical samples with lateral flow or fluorescence readout. For panels **D** and **E**: T = test band; C = control band.

### Evaluation of ADESSO performance on clinical samples in direct comparison to RT-qPCR and an antigen test

We used ADESSO to test a total of 195 clinical samples in direct comparison to RT-qPCR and an antigen test commonly used for the diagnosis of COVID-19^59^. To allow a fair comparison between the methods, we first selected 95 positive and 100 negative individuals (via COBAS RT-qPCR on NP swab). For each of these specimens, RNA was re-extracted and analyzed by both RT-qPCR (Tib Molbiol) and ADESSO using a lateral flow readout to simulate a POC test. Additionally, an antigen-based diagnostic test (RIDA QUICK SARS-CoV-2-Antigen) and ADESSO were performed directly on unextracted samples. Finally, we also obtained saline gargle specimens from the same individuals as an alternative sampling method (**Figure 2A**).

**Figure 2:**
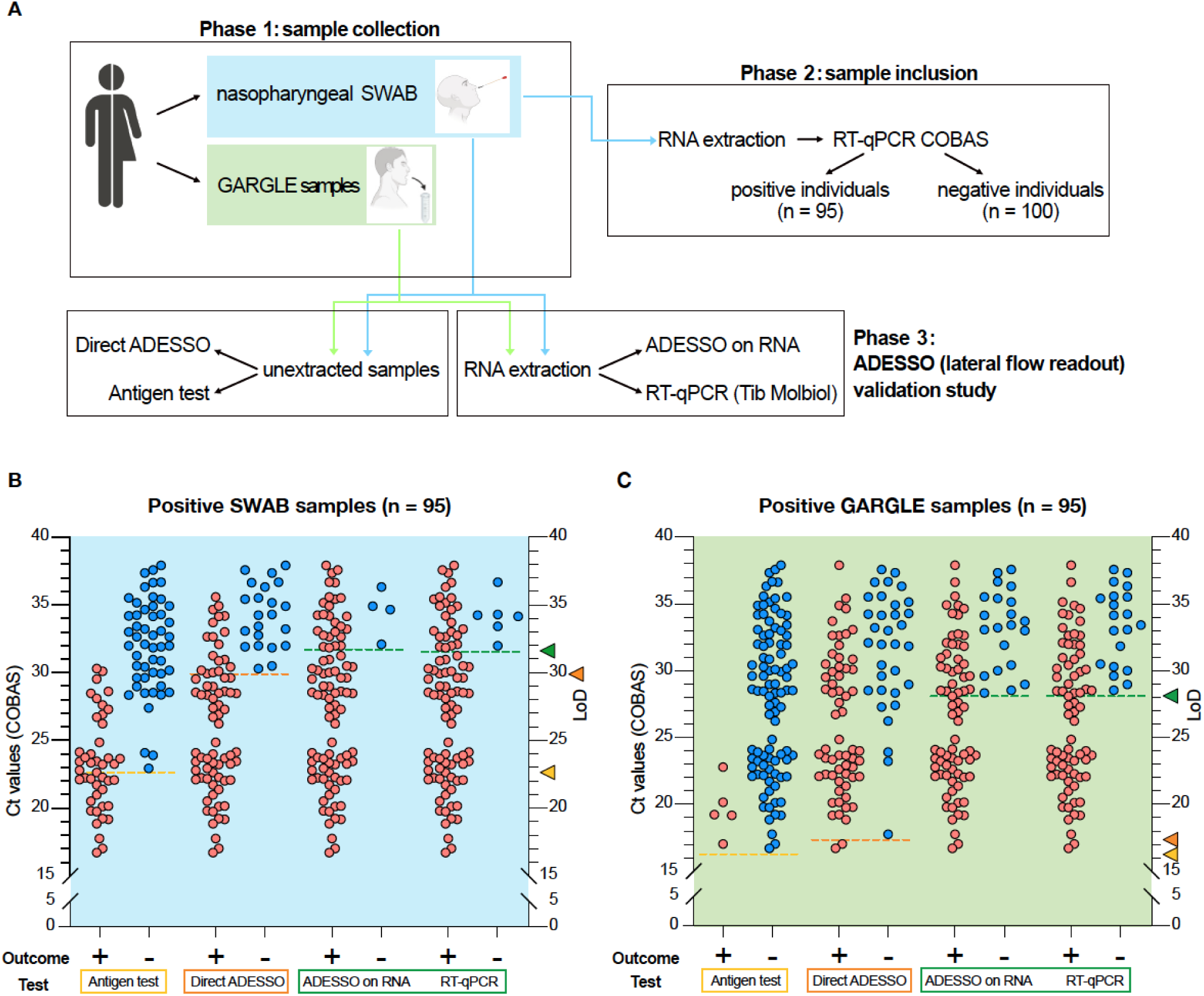
Evaluation of ADESSO performance on clinical samples in direct comparison to RT-qPCR and antigen test. **A**. Schematic of the validation study to assess ADESSO performance for SARS-CoV-2 detection in clinical samples in comparison with RT-qPCR (Tib Molbiol) and antigen test (RIDA QUICK SARS-CoV-2 Antigen). The COVID-19 status of the samples included in the study was initially determined by RT-qPCR (COBAS). ADESSO was performed on both extracted RNA and unextracted samples with lateral flow readout. **B** and **C**. SARS-CoV-2 detection and LoD evaluation in swab (**B**) and gargle (**C**) samples collected from 95 COVID-19 positive individuals using RIDA QUICK SARS-CoV-2 Antigen test, ADESSO and Tib Molbiol RT-qPCR. “Direct” ADESSO and RIDA QUICK SARS-CoV-2 Antigen test were performed on unextracted samples, while ADESSO and Tib Molbiol RT-qPCR were performed on extracted RNA. The LoD for each test is represented as a dotted line and the corresponding Ct value is indicated by an arrowhead on the right Y axis.

This randomly selected cohort of positive individuals covers the full distribution of viral titers between Ct 17 and Ct 38 (**Supplementary File 1**)^60^, thus allowing us to avoid bias during LoD evaluation due to sample size or viral load distribution^61^. The results of this experiment are summarized in **Table 1**. ADESSO on RNA extracted from swabs was able to correctly identify most positive samples (91 out of 95), resulting in a sensitivity of 96% and a LoD corresponding to Ct value ∼32 (**Figure 2B, S4A**). RT-qPCR (Tib Molbiol) performed on the same samples was largely in agreement with the COBAS RT-qPCR, with highly correlated Ct values (**Figure S4E**). Notably, using this method we were able to identify 89 out of 95 positive samples, resulting in a slightly lower sensitivity of 94% compared to ADESSO (**Table 1** and **Figure 2B, S4E**). Although ADESSO on unextracted swab samples resulted in a reduced sensitivity (77%) and LoD (Ct value 30), it still strikingly outperformed the antigen test, which only detected 44 out of 95 positive samples, resulting in a sensitivity of 46% and an LoD corresponding to Ct 23 (**Table 1** and **Figure 2B, S4B**). Additionally, while ADESSO and the RT-qPCR test showed 100% specificity, we observed one false-positive sample by antigen test (**Table 1**). Similar results were observed on saline gargle samples with a general drop in sensitivity and LoD for all the detection methods (**Table 1** and **Figure 2C**). Interestingly, this drop seems to be related to the sampling method. Indeed, higher RT-qPCR Ct values were observed in gargle specimens compared to their matched swab samples (**Figure S4E-G**). A similar drop in sensitivity was also reported in another study where a large cohort of paired nasopharyngeal swab-saliva samples was tested^62^.

**Table 1:** Positive and negative predictive values, sensitivity and specificity of ADESSO (with and without RNA extraction), Tib Molbiol RT-qPCR and RIDA QUICK SARS-CoV-2 antigen test on swab and gargle samples.

| Sampling method | Sample | Test | Test result | number |  |  | number/total number(percentage) |  |  |  |
| --- | --- | --- | --- | --- | --- | --- | --- | --- | --- | --- |
|  |  |  |  | Positive samples (N=95) | Negative samples (N=100) | Total samples (N=195) | Positive predictive value | Negative predictive value | Sensitivity | Specificity |
| SWAB | RNA | Tib Molbiol qRT-PCR | Positive | 89 | 0 | 89 | 89/89 (100%) |  | 89/95 (94%) |  |
|  |  |  | Negative | 6 | 100 | 106 |  | 100/106 (94%) |  | 100/100 (100%) |
|  |  | ADESSO | Positive | 91 | 0 | 91 | 91/91 (100%) |  | 91/95 (96%) |  |
|  |  |  | Negative | 4 | 100 | 104 |  | 100/104 (96%) |  | 100/100 (100%) |
|  | Lysate | ADESSO | Positive | 73 | 0 | 73 | 73/73 (100%) |  | 73/95 (77%) |  |
|  |  |  | Negative | 22 | 100 | 122 |  | 100/122 (82%) |  | 100/100 (100%) |
|  |  | RIDA QUICK antigen test | Positive | 44 | 1 | 45 | 44/45 (98%) |  | 44/95 (46%) |  |
|  |  |  | Negative | 51 | 99 | 150 |  | 100/150 (67%) |  | 99/100 (99%) |
| GARGLE WATER | RNA | Tib Molbiol qRT-PCR | Positive | 75 | 0 | 75 | 75/75 (100%) |  | 75/95 (79%) |  |
|  |  |  | Negative | 20 | 100 | 120 |  | 100/120 (83%) |  | 100/100 (100%) |
|  |  | ADESSO | Positive | 74 | 0 | 74 | 74/74 (100%) |  | 74/95 (78%) |  |
|  |  |  | Negative | 21 | 100 | 121 |  | 100/121 (83%) |  | 100/100 (100%) |
|  | Lysate | ADESSO | Positive | 62 | 0 | 62 | 62/62 (100%) |  | 62/95 (65%) |  |
|  |  |  | Negative | 33 | 100 | 133 |  | 100/133 (75%) |  | 100/100 (100%) |
|  |  | RIDA QUICK antigen test | Positive | 5 | 0 | 5 | 5/5 (100%) |  | 5/95 (5%) |  |
|  |  |  | Negative | 90 | 100 | 190 |  | 100/190 (53%) |  | 100/100 (100%) |

Altogether, ADESSO demonstrated similar sensitivity to RT-qPCR (Tib Molbiol) on extracted RNA and outperformed the antigen test when performed on unextracted samples. These results validate the high potential of ADESSO as a POC test for the detection of SARS-CoV-2 infected individuals.

### Adaptation of ADESSO for detection of SARS-CoV-2 variants: a flexible and powerful assay to rapidly identify specific variants or mutations

In order to demonstrate that ADESSO can be easily adapted for the detection of SARS-CoV-2 variants, we focused our attention on the two first known variants of concern: SARS-CoV-2 B.1.1.7 (Alpha) and SARS-CoV-2 B.1.351 (Beta). Both variants seem to have an enhanced transmissibility and might be more virulent^63–66^. Both Alpha and Beta variants contain several mutations in the spike gene, including deletions (e.g., ΔHV69-70 in Alpha) and mutation clusters (e.g., Δ242-244 and R246I in Beta) (**Figure 3A**). Finally, there is growing evidence that these new variants could impair the efficacy of current monoclonal antibody therapies and vaccines due to mutations in the spike gene^67–69^. To demonstrate the adaptability of ADESSO we modified our test to detect the deletion ΔHV69-70 specific to the B.1.1.7 strain. We called this adapted test ADESSO-Alpha (**Figure 3A**,**B**). First, we designed two crRNAs to distinguish between the Alpha variant (crRNA ΔHV69-70) or the corresponding original sequence of SARS-CoV-2 (crRNA HV69-70) (**Figure 3B**). We next tested RT-RPA primers designed against the region of SARS-CoV-2 genome containing HV69-70 and selected the more sensitive set for further analysis (**Figure S5A**). Finally, we performed a blind test on positive clinical samples carrying either the Alpha or Beta variant. Using ADESSO-Alpha (crRNA ΔHV69-70 and HV69-70) and we were able to correctly identify all the samples carrying the Alpha variant (samples #1-10, **Figure 3C**,**D** and **S5B**) and furthermore accurately distinguish these samples from those bearing the Beta strain (samples #11-13, **Figure 3C**,**D** and **S5B**). Using the standard ADESSO (crRNA/primers) we could detect all positive samples with the exception of sample #11 (**Figure 3E**). To assess this false negative, we sequenced the viral genome in the three samples carrying the Beta strain (#11-13) with the goal of determining whether additional uncharacterized mutations were occurring in the region targeted by ADESSO. This analysis revealed that indeed all the three samples shared a deletion (Δ242-244) within the central region bound by the forward primer used in the RT-RPA step of ADESSO (**Figure S5C**,**D**), demonstrating that ADESSO is resistant to deletions of several nucleotides within this region. Furthermore, sample #11 carried an additional mutation (R246I), within the region recognized by the 3’ end of the same primer (**Figure S5C**,**D**), which explains why this sample was only detected by ADESSO-Alpha and highlights the high specificity of the assay, which can discriminate even single-base changes.

**Figure 3:**
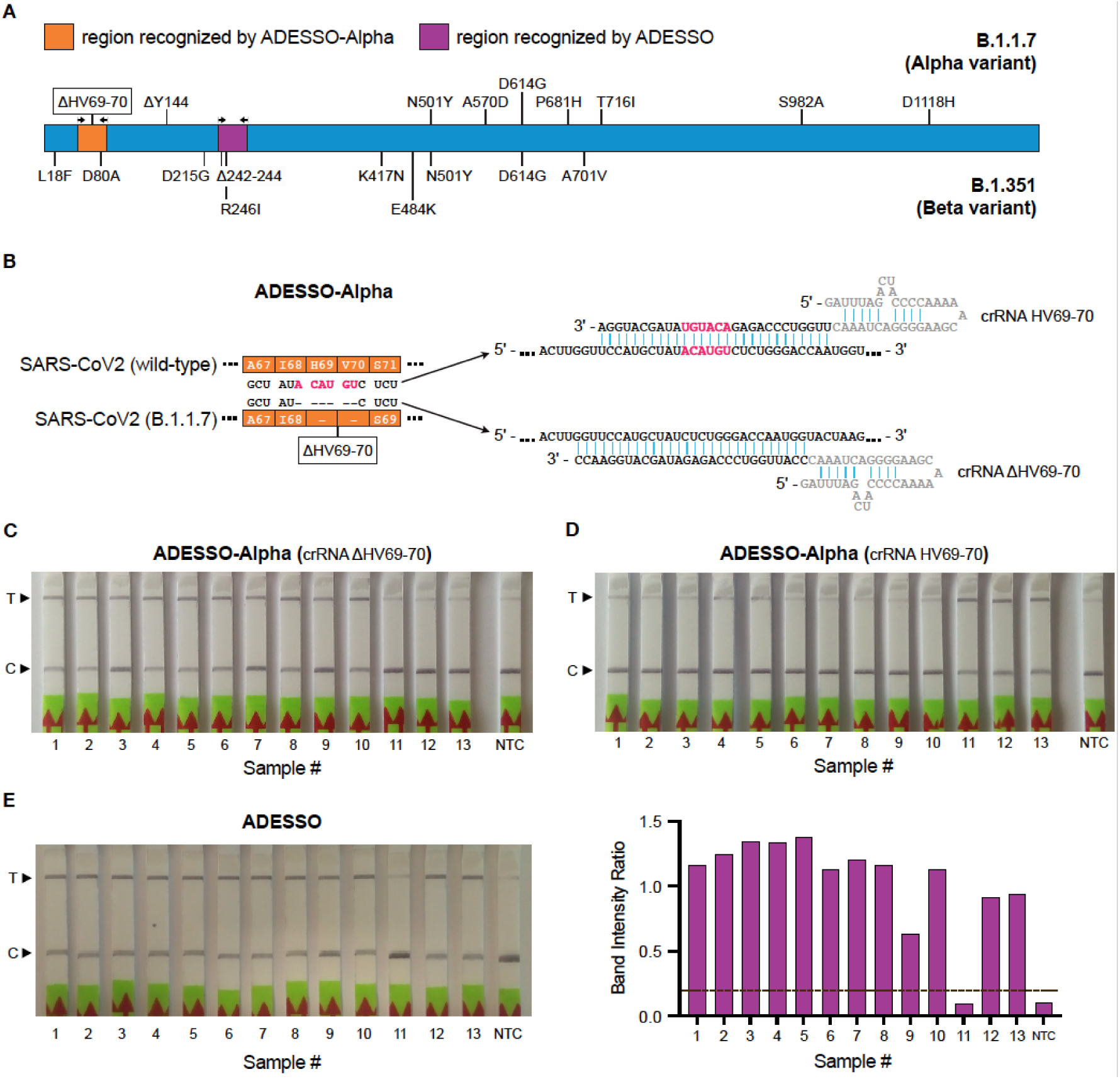
Adaptation of ADESSO for detection of SARS-CoV-2 variants: a flexible and powerful assay to rapidly identify specific variants or mutations. **A**. Schematic of SARS-CoV-2 S gene with annotation of the reported mutations for SARS-CoV-2 B.1.1.7 (Alpha, top) and B.1.351 (Beta, bottom) lineages. The regions of the S gene targeted by ADESSO and ADESSO-Alpha are indicated in purple and orange, respectively. **B**. Schematic of the S gene region containing the ΔHV69-70 deletion (highlighted in pink) specific for the B.1.1.7 variant in comparison with the original SARS-CoV-2 sequence and illustration of the binding of the specific crRNAs targeting the mutated (crRNA ΔHV69-70) or wild-type (crRNA HV69-70) sequence. The grey sequence in the crRNAs is called direct repeat (DR) and its stem-loop structure is needed for the recruitment of Cas13. **C**. SARS-CoV-2 B.1.1.7 variant detection by ADESSO-Alpha with ΔHV69-70 crRNA and (**D**) confirmation of the absence of the Alpha variant-specific deletion by ADESSO-Alpha with HV69-70 crRNA in 13 clinical samples carrying either the SARS-CoV-2 Alpha (B.1.1.7) or Beta (B.1.351) variant. **E**. Confirmation of the detection of SARS-CoV-2 by ADESSO in the same samples as in **C** and **D**. The band intensity ratios are shown in the bar plot on the right. For **C, D, E**: T = test band; C = control band; NTC = non template control.

Altogether, these results show how ADESSO can be readily adapted for the detection of SARS-CoV-2 variants of concern and even specific mutations. The entire adaptation of the test took less than one week, from the selection of a unique mutation for the Alpha variant to the validation of the adapted protocol, including designing and production of the specific reagents. This feature of our assay is a crucial aspect in the current phase of the COVID-19 pandemic, where quick and sequencing-independent detection of variants is essential to contain their spread.

## DISCUSSION

Weekly COVID-19 cases continue to rise in some countries despite multiple effective vaccines being distributed^1^. Effectively tracking new infections using the RT-qPCR-based COVID-19 diagnostic test is limited by the requirement for specific equipment, laboratory infrastructures and qualified personnel, often resulting in a long sample-to-result turnaround time. Despite widespread use, the lower sensitivity of antigen tests makes them suitable only for the detection of infected individuals with a high viral load^29^. CRISPR-Dx technologies are attractive for POC testing thanks to their programmability and independence of complex instruments. Here, we developed ADESSO, a Cas13a-based optimized method for highly sensitive COVID-19 testing. Overall, we tested 793 samples (393 positive and 400 negative, **Supplementary File 1**), in parallel comparison with RT-qPCR and antigen test. ADESSO has a sensitivity and specificity comparable to RT-qPCR when performed on RNA extracted from either swabs or gargle samples (**Table 1**). However, skipping the RNA extraction step allows more frequent testing, which is suggested by a recent model of viral dynamics to be essential for efficient identification and isolation of carriers to contain the pandemic^25^. To evaluate the potential of ADESSO as a POC test, we benchmarked its performance on unextracted swab samples against an antigen test, which represents the most commonly used POC diagnostic test at the moment. Remarkably, despite a decrease in sensitivity (77%), ADESSO largely outperformed the antigen test, which correctly detected less than half of the positive samples resulting in a sensitivity of 46% (**Table 1**). These results demonstrate that using ADESSO as a POC test on unextracted samples could considerably improve COVID-19 surveillance in comparison to the antigen tests currently in use.

The importance of testing is further highlighted by the constant emergence of worrisome SARS-CoV-2 variants, as India’s tragic crisis has shown^70^. This poses an additional challenge because mutations in the viral genome might impair both molecular and antigen-based tests, thus leading to false negative results. In these situations, being able to promptly adapt a test is fundamental and ADESSO offers such an advantage. In our study, in less than a week we adapted the test for the detection of the Alpha variant. Based on the publicly available SARS-CoV-2 sequences (https://www.gisaid.org), ADESSO can be adapted to any variant of concern, thus providing an all-in-one SARS-CoV-2 detection and variant identification tool prior to downstream sequencing. Even so, genomic surveillance to track the evolution of the virus remains of utmost importance for the discovery of new emerging and circulating variants^71^ and for providing the information needed to promptly adapt nucleic acid-based diagnostic tests such as ADESSO.

In order to understand what portion of the SARS-CoV-2 infected population ADESSO would be able to detect, we analyzed the Ct value distribution across a population of 6439 infected individuals among ambulatory patients presenting minimal to mild symptoms as well as asymptomatic people who had contact with COVID positive individuals between October 1st, 2020 and July 31st, 2021. We observed a distribution ranging from Ct 17 to Ct 38. This observation confirms the fact that also these individuals can manifest high viral loads and therefore be infectious^72,73^, thus facilitating oblivious spread of the virus. Based on the LoD evaluation shown in this study, by applying ADESSO on unextracted swab samples an estimated ∼70% of the infected population would be successfully detected (**Figure 4**). This portion is remarkably higher (31% more) than the one detected by antigen tests that are currently commonly used in our society. Notably, mathematical models show that successful identification and isolation of 50% of infected individuals is already sufficient to flatten the infection curve^74^. Our test exceeds this fraction in all conditions (**Table 1** and **Figure 4**), strongly suggesting that immediate and widespread application of ADESSO would be of great help to contain the pandemic. The portion of infected individuals potentially detectable by ADESSO might also increase in the case of infection by the Delta variant, for which a 1000x higher average viral load was observed^75–77^.

**Figure 4:**
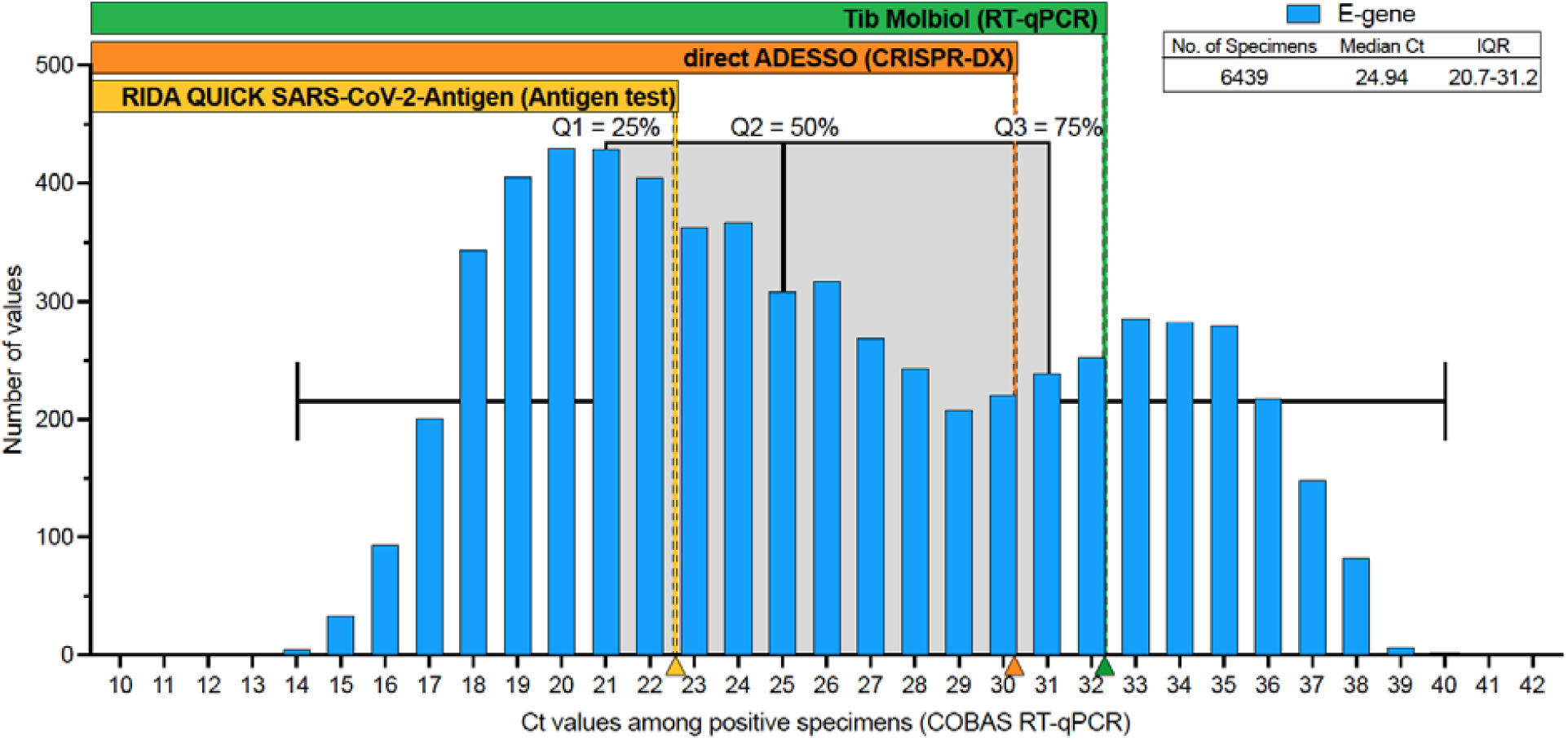
Ct value distribution across a population of non- or mildly symptomatic COVID-19 positive individuals. The histogram shows the distribution of Ct values detected by using COBAS RT-qPCR (SARS-CoV-2 E gene) across 6439 positive samples. These were collected from ambulatory patients showing minimal to mild symptoms. Median and interquartile range (IQR) of the distribution are indicated in the box at the top-right and illustrated by the box plot in gray behind the histogram. Quartiles (Q) 1, 2, 3 are also indicated to highlight the 25%, 50% and 75% portions of the population, respectively. Arrowheads on the x axis represent the Ct values corresponding to the LoD of the three different detection methods, as evaluated in this work (Figure 2). The rectangles above the histogram indicate the range of Ct values and the portions of specimens that would be detected by the three tests compared in this study (antigen test in yellow, direct ADESSO in orange and RT-qPCR in green).

Furthermore, our results show a disagreement between LoD on serial dilutions of synthetic viral genome and LoD in clinical samples. Despite the “synthetic” LoD of 2.5 cp/μl (∼Ct 35; **Figure 1**), the real clinical sensitivity of ADESSO corresponds to Ct 30-32 (**Figure 2**) and the same is true for other studies although it has never been clearly articulated^44,47,54^. This aspect highlights that an extensive validation on real clinical samples covering the full range of viral titers, as the one shown here, is necessary to determine the real LoD of a diagnostic test. This is crucial to allow a fair comparison between sensitivities resulting from independent studies, which can be greatly influenced by the choice of the tested population. To our knowledge, it is the first time that such an extensive study on clinical samples in direct comparison to both RT-qPCR and antigen test has been reported for CRISPR-Dx technologies.

Finally, we calculated a cost per reaction of 2.64€ and 4.82€ for fluorometric and lateral-flow detection, respectively (**Supplementary File 2**), which is comparable to antigen tests and lower than any other detection method (**Table 2**). Altogether, ADESSO is cheaper than any RT-qPCR-based COVID-19 diagnostic test^78^ and offers a more accessible option for widespread and more frequent testing. At the same time, ADESSO is comparable to the commonly used antigen tests in terms of cost^79^, but offers a higher sensitivity.

**Table 2:** Comparison of ADESSO assay with other widespread assays for the detection of SARS-CoV-2.

|  | Antigen test | ADESSO |  | Rapid RT-qPCR | RT-qPCR |
| --- | --- | --- | --- | --- | --- |
|  |  | Direct | on RNA |  |  |
| Clinical LoD (Ct value) | 22<sup>(30)</sup> - 25<sup>(28)</sup> | 30 | 32 | 29<sup>(27)</sup> - 35<sup>(28)</sup> | 36<sup>(26)</sup> |
| Assay reaction time | 5-20 min | 60 min | 60 min * | 45 min | 120 min * |
| RNA extraction | No | No | Yes | No | Yes |
| Sophisticated equipment needed | No | No | No | Yes | Yes |
| POC suitability | Yes | Yes | No | Yes | No |
| Cost per reaction | 4,80€<sup>(79)</sup> | 2-5€ | 2-5€ * | 15-20€<sup>(78)</sup> | 9,11€<sup>(79)</sup> |
| * without RNA extraction |  |  |  |  |  |

With the COVID-19 pandemic now deep into its second year, it has become clear that time plays a critical role in the management of such an emergency. In order to control it, while waiting to evaluate the long-term effect of the ongoing worldwide vaccination campaign, rapid detection of new infections, quick tracing of contacts and prompt reaction to emerging new variants are key-factors. Slowly but undeniably, the race against SARS-CoV-2 has turned from a sprint into a marathon. If we want to keep up, we need to take action faster than the virus evolves. The time is now for ADESSO to join the race.

## Data Availability

All the data will be available after the publication of this article.

## SUPPLEMENTARY FIGURES

**Figure S1:**
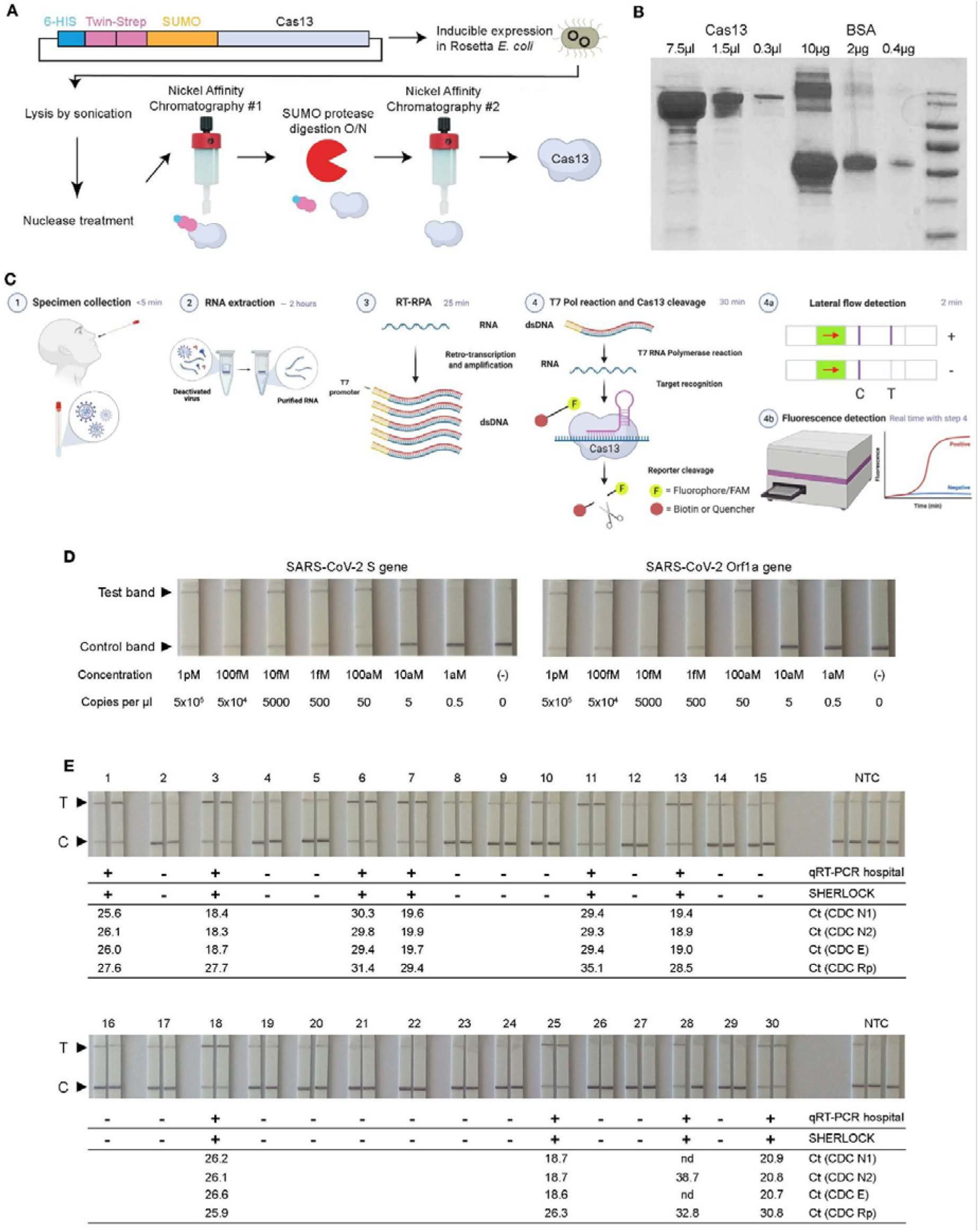
Generation of LwaCas13a and first attempt of SHERLOCK. **A**. LwaCas13a protein purification. The LwaCas13 fusion construct also encodes multiple affinity tags and a protease recognition site at the N terminus of the polypeptide. We have utilized the 6xHIS tag as the basis for our relatively inexpensive purification, while others have developed an alternative protocol based on the Strep-tags^57^. After expression in Rosetta cells (inducible via the Lac operon), the cells are lysed by sonication and the nucleic acid contained within the lysate is digested. The fusion protein is then purified by nickel-affinity chromatography. The purified fusion protein is digested with SUMO protease, which cleaves the tags and majority of the SUMO site off of the mature protein. The SUMO protease and in-tact affinity tags are then removed from the sample by re-applying the sample to the nickel column, leaving >98% pure Cas13. We also employ a size exclusion chromatography step to remove any aggregated Cas13 protein (not pictured). **B**. Serially diluted amounts of pure Cas13 were analyzed by coomassie staining after conventional SDS-PAGE, revealing a prominent band at the appropriate molecular weight and only minor contaminants. A serial dilution of BSA was also run as an estimate of protein concentration by densitometry (which was also validated by BCA assay). **C**. Graphic of SHERLOCK experimental workflow to detect SARS-CoV-2 in RNA extracted from clinical samples with lateral flow readout. **D**. SHERLOCK sensitivity on serial dilutions of an IVT fragment of SARS-CoV-2 S and Orf1a genes. **E**. Comparison of SARS-CoV-2 detection on RNA extracted from 30 clinical samples via SHERLOCK and RT-qPCR (Medical University Hospital Mannheim (RT-qPCR hospital) or CDC 2019-nCoV Real-Time RT-PCR Diagnostic Panel). SHERLOCK was performed on SARS-CoV-2 S gene; RT-qPCR at the Medical University Hospital Mannheim was performed on SARS-CoV-2 E and Orf1a genes; CDC RT-qPCR was performed on SARS-CoV-2 N1, N2 and E genes (CDC N1, N2, E) and human RNase P (CDC Rp) as RNA quality control. T = test band; C = control band; nd = not detected; NTC = non template control.

**Figure S2:**
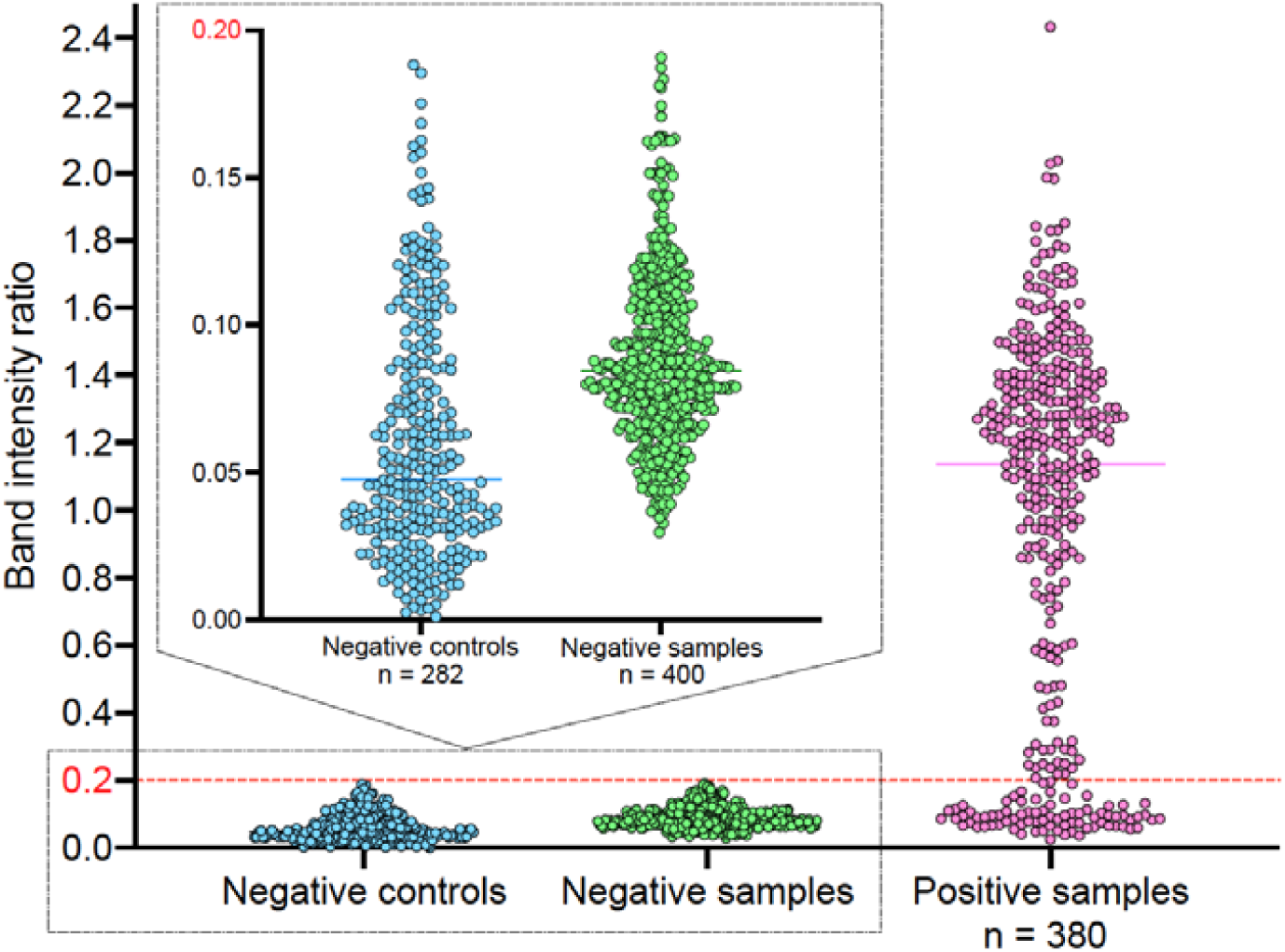
SARS-CoV-2 clinical samples. Definition of threshold between positive and negative results. The bar plot shows the band intensity ratios of all the negative controls utilised in this study (282, in blue) together with the negative (100*4 = 400, in green) and positive (95*4 = 380, in pink) clinical samples analysed in Figure 2. These data were used to define a threshold band intensity ratio of 0.2 to distinguish between positive and negative samples.

**Figure S3:**
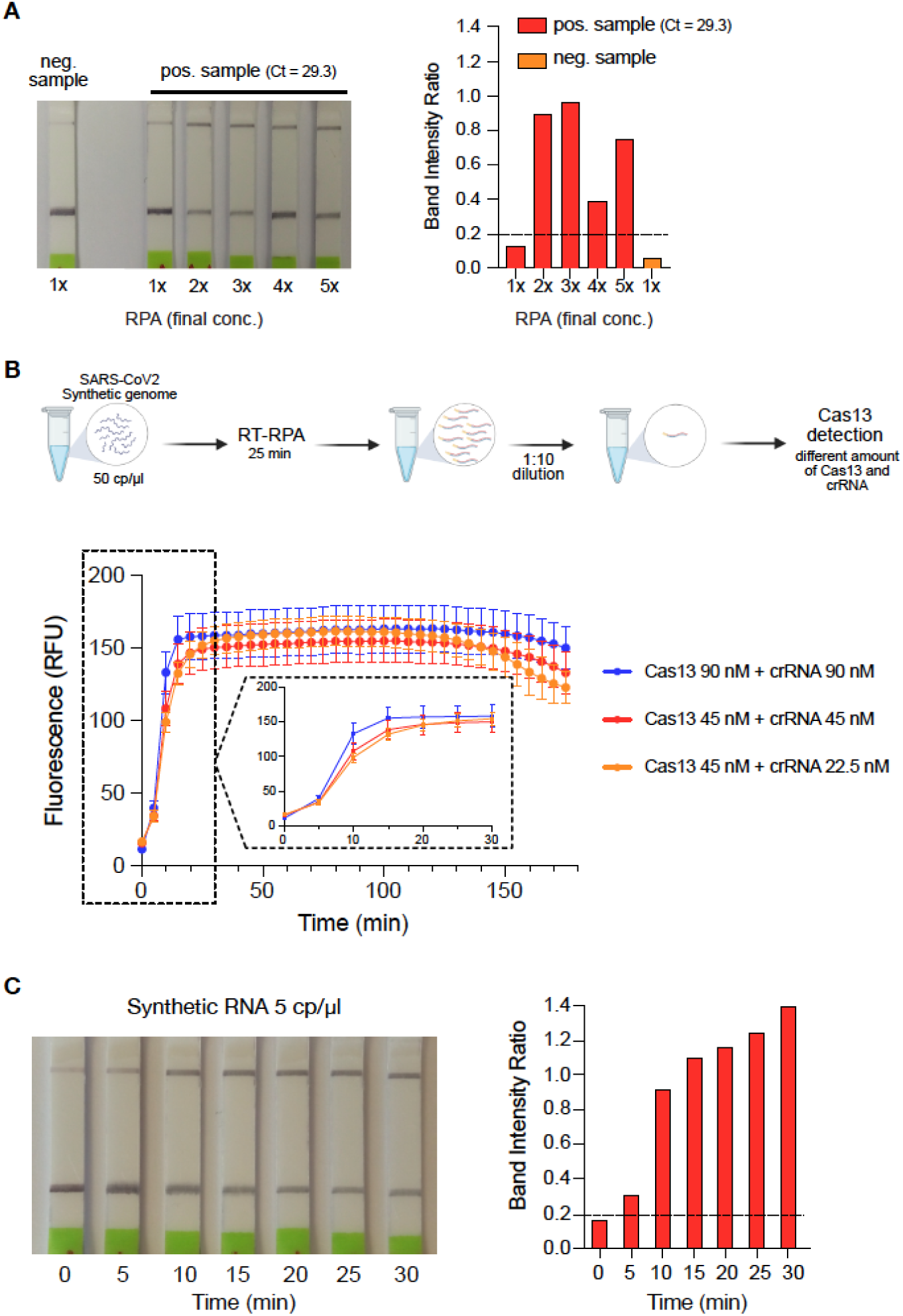
ADESSO: an optimised and highly sensitive SHERLOCK assay. **A**. Optimisation of direct SHERLOCK sensitivity with lateral flow readout by increasing the RPA reagents to detect a false negative sample (Sample #11, Supplementary File 1). A true negative sample (Sample #10, Supplementary File 1) is used as negative control. **B**. Scheme of the experiment for the optimisation of the Cas13 reaction kinetics with fluorescence readout using different amounts of Cas13 protein and crRNA and complete 3-hour analysis of the fluorescence signal at different time-points. The enlarged window shows the first 30 minutes of the reaction. **C**. Time-point analysis of the Cas13 reaction to determine the shortest incubation time required to detect a positive signal with lateral flow readout.

**Figure S4:**
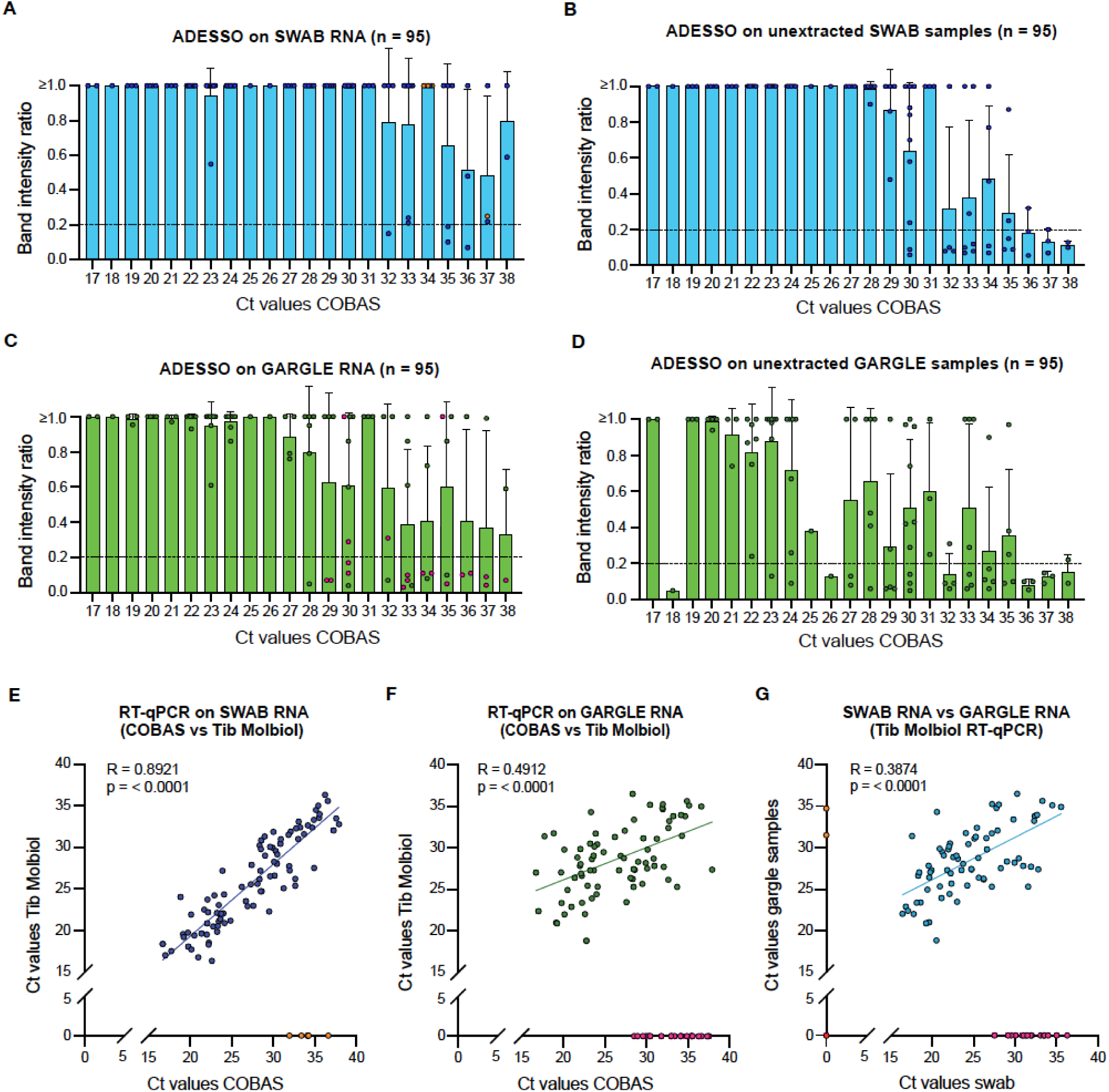
Evaluation of ADESSO performance on clinical samples in direct comparison to RT-qPCR. **A**. ADESSO performance on RNA extracted from swab specimens in comparison with COBAS RT-qPCR. Negative samples by Tib Molbiol RT-qPCR are represented in orange. **B**. ADESSO performance on unextracted swab specimens in comparison with COBAS RT-qPCR (performed on RNA extracted from swabs). **C**. ADESSO performance on RNA extracted from gargle samples in comparison with COBAS RT-qPCR (performed on RNA extracted from swabs). Negative samples by Tib Molbiol RT-qPCR are represented in pink. **D**. ADESSO performance on unextracted gargle samples in comparison with COBAS RT-qPCR (performed on RNA extracted from swabs). **E**. Correlation analysis of Ct values obtained with Tib Molbiol RT-qPCR (y axis) or COBAS RT-qPCR (x axis) on RNA extracted from swab specimens. Negative samples by Tib Molbiol RT-qPCR are represented in orange and are excluded in the calculation of the correlation (R). **F**. Correlation analysis of Ct values obtained with Tib Molbiol RT-qPCR (y axis) and COBAS RT-qPCR (x axis) on RNA extracted from gargle samples. Negative samples by Tib Molbiol RT-qPCR are represented in pink and are excluded in the calculation of the correlation (R). **G**. Correlation analysis of Ct values obtained after Tib Molbiol RT-qPCR on RNA extracted from gargle (y axis) and swab (x axis) samples. Negative swab samples are represented in orange; negative gargle samples are represented in pink; negative samples both as gargle and swab are represented in red. All the negative samples are excluded in the calculation of the correlation (R). In panels **A, B, C, D** only the band intensity ratios of the positive samples are shown (n = 95). Values higher than 1 are plotted as equal to 1 for better visualisation. LoD = Limit of detection. Ct values are rounded to the nearest whole number for the sake of visualisation. The exact Ct values are listed in Supplementary File 1.

**Figure S5:**
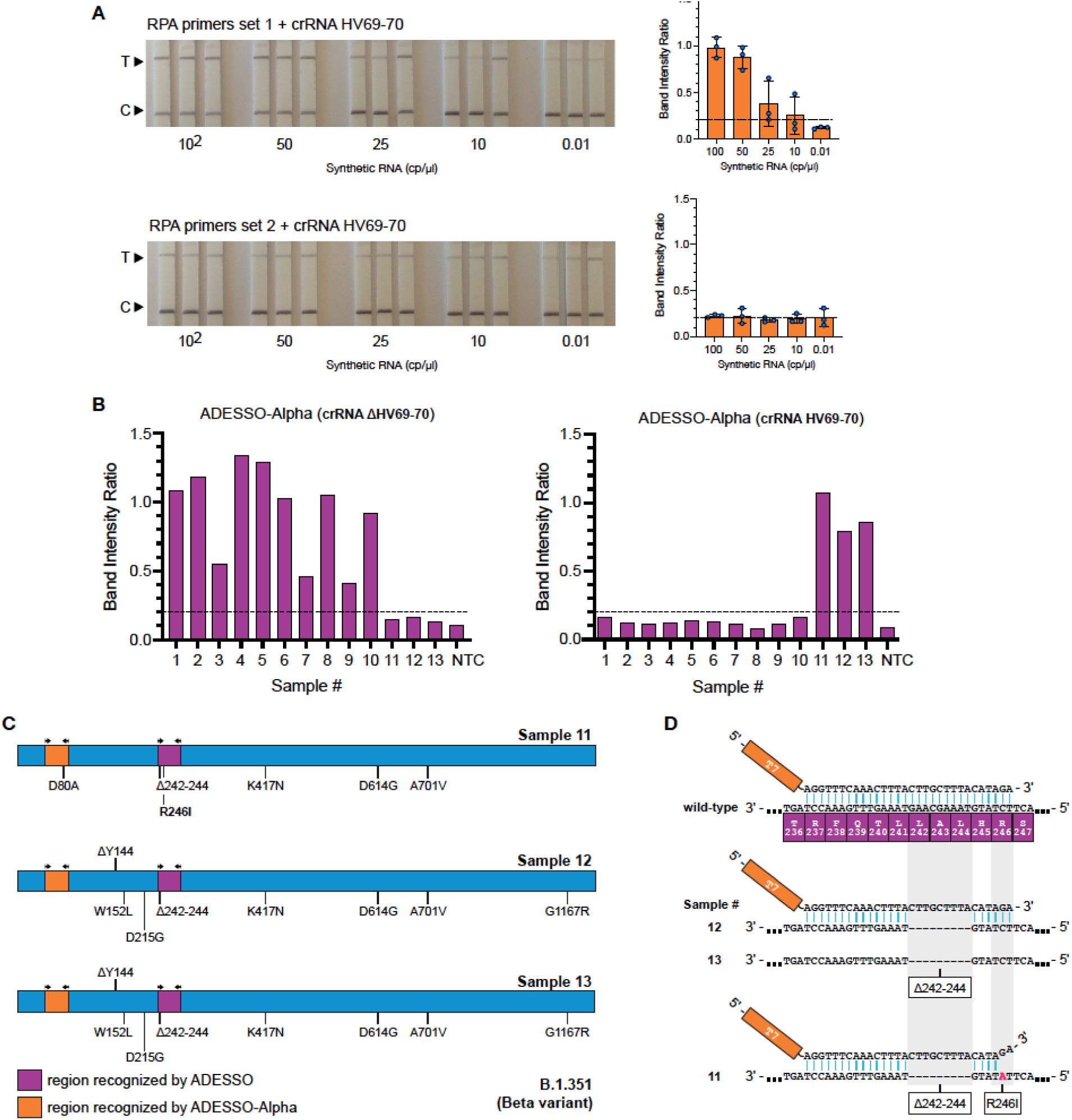
Adaptation of ADESSO for detection of SARS-CoV-2 variants. **A**. RPA primers optimization to amplify the region of SARS-CoV-2 S gene surrounding the B.1.1.7 variant-specific deletion causing ΔHV69-70. Two combinations of the same forward primer with two alternative reverse primers were tested (set 1 and set 2) on serial dilutions of SARS-CoV-2 synthetic genome (wild-type sequence). Cas13 detection was performed using crRNA HV69-70. Band intensity ratios are shown on the right side. T = test band; C = control band. **B**. Band intensity ratios of 13 clinical samples carrying either the Alpha (B.1.1.7) or Beta (B.1.351) SARS-CoV-2 variant tested by ADESSO-Alpha. The corresponding lateral flow strips are shown in Figure 3D and E. The bar plot on the left illustrates the results of the SARS-CoV-2 B.1.1.7 variant detection by ADESSO-Alpha with crRNA ΔHV69-70. The bar plot on the right illustrates the results of the confirmation of the presence of SARS-CoV-2 B.1.351 variant by ADESSO-Alpha with crRNA HV69-70. NTC = non template control. **C**. Schematic of SARS-CoV-2 S gene with annotation of the mutations identified in three patients (clinical samples #11, #12 and #13) carrying the Beta variant. The regions of the S gene targeted by ADESSO and ADESSO-Alpha are indicated in purple and orange, respectively. **D**. Schematic of the binding of the forward RPA primer used in ADESSO to the complementary region in the original SARS-CoV-2 sequence (top), in clinical samples #12 and #13 carrying the Beta variant with the deletion Δ242-244 (middle) and in sample #11 carrying the Beta variant with an additional mutation (R246I) (bottom). The positions of the Δ242-244 deletion and the R246I mutation are highlighted in grey. The point mutation causing the R246I substitution is marked in red.

## METHODS

### Protocols

The RT-RPA and Cas13 reaction protocols used for each experiment are provided in Supplementary File 2 with reference to the corresponding figures. The exact volumes are given for one single reaction.

### Reagents and materials

Detailed information about reagents and material used in this study is provided in Supplementary Table 3.

### Cas13 purification

Plasmid encoding LwaCas13 (pC013 - Twinstrep-SUMO-huLwCas13a was a gift from Feng Zhang (Addgene plasmid # 90097 ; http://n2t.net/addgene:90097 ; RRID:Addgene_90097)^40^ was transformed into Rosetta cells and purified according to established protocols with substantial modification. Single colonies were inoculated into 25 ml Terrific Broth (TB) (100 μg/ml AMP) and grown to an OD of 0.6 at 37°C degrees while shaking at 150 rpm. The suspension was chilled for 30 min at 4°C and subsequently induced with 0.5 mM IPTG and left shaking for an additional 16h at 21°C. Cells were harvested by centrifugation at 5 k rpm for 15 min at 4°C. The pellet was resuspended in 4x (wt/vol) supplemented lysis buffer (12 cOmplete Ultra EDTA-free tablets, 600 mg of lysozyme and 6 μl of benzoase to lysis buffer (20 mM Tris pH 8.0, 500 mM NaCl, 1 mM DTT)) and lysed by sonication. Lysate was cleared by centrifugation at 10 k rpm for 1h at 4°C. Supernatant was purified using a 1 ml HIS-Trap column (Cytiva) slurry and affinity chromatography was performed using the ÄKTA pure system with lysis buffer for washing steps and an imidazole gradient for elution. After initial purification, the protein sample was incubated with SUMO protease (ThermoScientific) as per the manufacturer’s instructions at 4°C overnight to remove the affinity tags. The sample was then re-applied to a 1 ml HIS-Trap column. Both the SUMO protease (which itself has a 6xHIS tag) and the cleaved affinity tag bind to the resin, while pure Cas13 eluted in the wash step. A final size-exclusion chromatography step was performed using the ÄKTA pure system using 10 mM HEPES pH 7.0, 5 mM MgCl2, 1 M NaCl and 2 mM DTT as gel filtration buffer on a Superdex 16/600 column.

### Synthetic SARS-CoV-2 RNA

Fully synthetic SARS-CoV-2 RNA was purchased from Twist Biosciences (MT007544.1 or MN908947.3). In order to test SHERLOCK sensitivity, serial dilutions were prepared in water or in saline, from the initial concentration of 10^6^ cp/μl to 0.01 cp/μl.

### Synthetic SARS-CoV-2 S gene and Orf1ab gene RNA fragments

SARS-CoV-2 RNA, a kind gift of Prof. Bartenschlager (DKFZ, Heidelberg), was used for OneStep RT-PCR (Qiagen, #210212) as follows: 11 μl of nuclease-free water, 5 μl of 5x OneStep RT-PCR buffer, 1 μl of dNTP mix (10mM each), 1.5 μl of each primer (forward and reverse, both 10μM) and 1 μl of OneStep RT-PCR Enzyme Mix were added to 4 μl of denatured RNA. The primers used for the amplification of SARS-CoV-2 S gene and Orf1a gene are listed in Supplementary Table 3. The RT-PCR protocol was run as follows: retrotranscription at 50°C for 30 min, denaturation at 95°C for 15 min, followed by 40 cycles of denaturation at 94°C for 30 sec, annealing at 61°C (Orf1a gene) or 62°C (S gene) for 30 sec and elongation at 72°C for 5 sec. In the end a final elongation step at 72°C was run for 10 min. PCR clean-up was performed on the RT-PCR products according to the manufacturer’s instructions (Macherey-Nagel, #740609.250). The purified DNA was in-vitro-transcribed into RNA with the HiScribe T7 Quick High Yield RNA Synthesis Kit (NEB, #E2050S) following the suggested protocol for short transcripts. The IVT products were then treated with DNase I (HiScribe T7 Quick High Yield RNA Synthesis Kit, NEB, #E2050S) and purified with Monarch RNA Cleanup Kit (NEB, #T2050). The concentration of the purified products was determined by Nanodrop and Qubit. In order to test SHERLOCK sensitivity, serial dilutions were made in water from a concentration of 1μM to 1aM.

### Human clinical specimen collection

Clinical specimens were collected at the Medical University Mannheim, Germany. The Ethics Committee II of the University of Heidelberg (Medical Faculty of Mannheim) ruled the ethics for all the clinical samples used in this study. The committee reviewed and approved the proposal for the collection and use of NP and gargle samples (ref. 2020-556N). NP swabs and gargle samples were collected from ambulatory patients presenting minimal to mild symptoms or sent by the German Health Department after having contact with a SARS-CoV-2 positive person. After verbal and visual instruction gargling was performed with 8 ml of sterile 0,9% saline (Fa. Fresenius Kabi, Bad Homburg, Germany). Samples were collected in sterile containers without additives and stored at 4°C until testing with PCR within 36 h. NP specimens were collected with flocked swabs (Improswab, Fa. Improve Medical Instruments, Guanzhou/China) and washed out with 2 ml 0,9% saline within 12 h of collection. For sample inclusion in the validation study and side-by-side comparison of ADESSO and RT-qPCR, initial PCR was performed on NP swab samples as part of routine clinical care using the cobas 6800 system (Roche, Penzberg, Germany) according to the manufacturer’s instructions. Based on the results of the initial PCR, 95 positive and 100 negative samples were selected.

### RNA extraction

For the first blind test (Figure S1), RNA was extracted from the clinical samples with the QIAamp® Viral RNA Mini kit (Qiagen, #52904) following the manufacturer’s instructions (140μl of swab were extracted and eluted in 60μl). For the validation study (Figure 2), RNA was extracted from 200 µl of the selected gargle and NP specimens with the MagnaPure Compact System (Roche, Penzberg, Germany) using the Nucleic Acid isolation Kit I (Roche) resulting in 100 µl of eluate. Residual volume of gargle and NP specimens was stored at 4°C and sent to the DKFZ for further analysis.

### RT-qPCR

CDC taqman RT-qPCR initially (Figure S1) was performed in technical triplicates according to published protocols^80^, which we adapted to a 384-well plate format and a reduced reaction volume of 12.5 µl. The reaction was performed using the Superscript III One-Step RT-PCR kit with Platinum Taq Polymerase. Magnesium sulphate and BSA were added to the reaction to a final concentration of 0.8 mM and 0.04 μg/μl, respectively. Primers and probes for the viral N1 and N2 and the human RNase P genes were added as ready-made mix (1 µl; Integrated DNA Technologies Belgium; CatNo. 10006713). The E-gene probes and primers (GATC, Germany) were used at final concentrations of 500 nM for each primer and 125 nM for the probe. ROX was added to a final concentration of 50 nM. PCR was performed in a QuantStudito 5 thermocycler, with cycling conditions 55°C for 10 min, 95°C for 3 min, followed by 45 cycles of 95°C for 15 s and 58°C for 30 s.

For the validation study (Figure 2), real-time PCR of 10 µl RNA-eluate was performed on a BioRad CX96 cycler using the Sarbeco E-Gen-Kit (Fa. Tib Molbiol, Berlin, Germany) following the manufacturer’s instructions. Residual volume of extracted RNA from gargle and NP specimens was stored at -20°C and sent to the DKFZ for further analysis.

### Lysis of clinical samples for direct SARS-CoV-2 detection

Clinical samples were lysed for direct ADESSO assay (Figure 2) as follows: after vortexing, 10μl of sample were mixed with 10μl of QuickExtract DNA Extraction solution (Lucigen, #QE09050) enriched with Rnase Inhibitor, Murine (NEB, #M0314) at a final concentration of 4U/μl. Samples were then incubated at 95°C for 5 min. After incubation, samples were mixed by vortexing and spun down for 15 seconds at 10.000g. Finally, 5.6 μl of sample (for RT-RPA 2X) were collected from the upper liquid phase, carefully avoiding to aspirate any precipitate, and used in the RT-RPA step.

### crRNA synthesis and purification

CRISPR-RNAs (crRNAs) were either designed in our lab or synthesised by Integrated DNA Technologies (IDT). All crRNAs used in this study are listed in Supplementary Table 3. To produce the crRNAs in our lab we followed a previously published protocol^57^. In short, the templates for the crRNAs were ordered as DNA oligonucleotides from Sigma-Aldrich with an appended T7 promoter sequence. These oligos were annealed with a T7-3G oligonucleotide, and used in an in vitro transcription (IVT) reaction (HiScribe T7 Quick High Yield RNA Synthesis Kit, NEB, #E2050S). The crRNAs were then purified using Agencourt RNAClean XP Kit (Beckman Coulter, #A63987). The correct size of the crRNAs was confirmed on a UREA gel and the concentration evaluated by nanodrop. Aliquots of 10ng/μl of each crRNA were produced to avoid repeated freeze and thaw cycles and stored at -80°C.

### Reverse Transcriptase Recombinase polymerase amplification (RT-RPA)

RT-RPA reactions were carried out with TwistAmp Basic (TwistDx, #TABAS03KIT) with the addition of M-MuLV Reverse Transcriptase (NEB, #M0253) and RNase Inhibitor, Murine (NEB, #M0314). Reactions were run at 42°C for 45 minutes in a heat block. Here are the details for the optimised reaction (so called RT-RPA 2X): two lyophilized pellets TwistAmp Basic are used to prepare the following master mix for 5 reactions: 59 μl of Rehydration Buffer (RB) are mixed with 2,5 μl of each primer (forward and reverse) at a concentration of 20μM, 1.5 μl of M-MuLV RetroTranscriptase (200U/μl - NEB, #M0253) and 1,5 μl of Rnase Inhibitor, Murine (40U/μl - NEB, #M0314). The RB-primer-enzyme mix is used to rehydrate two pellets and finally 5μl of MgOAc are added. The complete mix is aliquoted (14.4μl) on top of 5,6 μl of each sample. The RT-RPA protocol was optimised throughout the study. To avoid any confusion, we provide an additional file (Supplementary File 2) with detailed protocols for each experiment presented in this work. All RPA primers used in this study are listed in Supplementary Table 3 and were designed following the provided guidelines^57^.

### Cas13 cleavage reaction for lateral flow readout

The optimised reaction mix for Cas13 activity was prepared by combining 4.3 μl of nuclease-free water, 1 μl of cleavage buffer (400mM Tris pH 7.4), 1 μl of LwaCas13a protein diluted in Storage Buffer (SB)^57^ to a concentration of 126.6 μg/ml, 0.5 μl of crRNA (40 ng/μl), 0.5 μl of lateral flow reporter (IDT, diluted in water to 20 μM), 0.5 μl of SUPERase-In RNase inhibitor (ThermoFisher Scientific, #AM2694), 0.4 μl of rNTP solution mix (25mM each, NEB, #N0466), 0.3 μl of NxGen T7 RNA Polymerase (Lucigen, #30223-2) and 0.5 μl of MgCl_2_ (120mM). 1 μl of the RT-RPA-amplified product was then added to the mix and, after vortexing and spinning down, the mixture was incubated for 10 minutes at 37°C in a heat block. The Cas13 protocol was optimised throughout the study. To avoid any confusion, we provide an additional file (Supplementary File 2) with detailed protocols for each experiment presented in this work.

### Lateral flow readout

Lateral flow detection was performed using commercially available detection strips (Milenia HybriDetect 1, TwistDx, Gießen, #MILENIA01). The 10μl-LwaCas13a reactions were transferred to a tube already containing 80 μl of HybriDetect Assay buffer. After vortexing and spinning down the reaction mix, a lateral flow dipstick was added to the reaction tube. The result was clearly readable after one minute. Once the whole reaction volume was absorbed, the dipstick was removed and photographed with a smartphone camera for band intensity quantification performed with the freely available ImageJ image processing program^81^. The results are shown as intensity ratio (test band/control band) and test were considered positive for value of intensity ratio above 0.2 based on the results shown in Figure S2.

### Cas13 cleavage reaction for fluorescence readout

The reaction mix for Cas13 activity was prepared by combining 8.6 μl of nuclease-free water, 2 μl of cleavage buffer (400mM Tris pH 7.4), 2 μl of LwaCas13a protein diluted in Storage Buffer (SB) to a concentration of 126.6 μg/ml, 1 μl of crRNA (40ng/μl), 1 μl of fluorescent reporter (IDT, diluted in water to a final concentration of 4 μM), 1 μl of RNase inhibitor, Murine (NEB, #M0314), 0.8 μl of rNTP solution mix (25mM each, NEB, #N0466), 0.6 μl of NxGen T7 RNA Polymerase (Lucigen, #30223-2) and 1 μl of MgCl2 (120mM). 2 μl of the RT-RPA-amplified product was then added to the mix. The 20μl-LwaCas13a reactions were transferred in 5μl-replicates (4 wells each sample) to a 384-well, round, black-well, clear-bottom plate (Corning, #3544). The plate was briefly spun down at 500g for 15 sec to remove potential bubbles and placed into a pre-heated GloMax® Explorer plate reader (Promega) at 37°C.

### Fluorescence readout

Fluorescence was measured every 5 min for 3 h. Data analysis, if not otherwise stated, was performed at the 30-min time-point.

### RNAse activity detection assay

In order to check for RNase activity in clinical samples, 10 μl of a negative swab sample (Figure 1A) were mixed with 10 μl of QuickExtract DNA Extraction Solution with or without RNase Inhibitor, Murine (NEB, M0314) at a final concentration of 4U/μl. The samples were then incubated at 95°C for 5 min. After incubation, RNaseAlert substrate v2 (RNaseAlert Lab Test Kit v2, #4479768, Thermo Fisher Scientific) was added at a final concentration of 200nM. The samples were mixed by vortexing, spun down and incubated at RT for 30 min in the dark. After incubation, the samples were transferred to a 384-well, round, black-well, clear-bottom plate (Corning, #3544) in 5μl-replicates (4 wells each sample). The plate was briefly spun down at 500g for 15 sec to remove potential bubbles and placed into a GloMax Explorer plate reader (Promega). RNaseAlert substrate fluorescence was measured every 5 min for 30 min. Data analysis, if not differently stated, was performed at the 5-min time-point.

### Antigen test

For the validation study (Figure 2), RIDA QUICK SARS-CoV-2 Antigen test was performed following the manufacturer’s instructions^82^.

## ACKNOWLEDGEMENTS

We thank the transport service staff, German Cancer Research Center (DKFZ), for providing excellent transportation services of all the clinical samples from the Medical University Mannheim to the DKFZ. The work was supported by a grant from the Ministry of Science, Research and the Arts of Baden-Württemberg for COVID-19 research (grant agreement no. Kap. 1499 TG 93 to Dr. Riccardo Pecori, Prof. Dr. Nina Papavasiliou (DKFZ) and Prof. Dr. Thomas Miethke (Medical Faculty of Mannheim)). Several schematics presented here were created with Biorender.com.

## AUTHOR CONTRIBUTIONS

BC, RP, and FNP designed the experiments. JPV and AH produced Cas13 protein. BC and RP performed all the experiments using SHERLOCK/ADESSO. PB, KHM, MS, PG and BR optimized and performed confirmative RT-qPCR on clinical samples using CDC protocol. MK collected the specimens and AGK performed the RT-qPCR on clinical samples using Tib Molbiol under the supervision of SW. BM analysed the RT-qPCR data. SA quantified the bands of the lateral flow strips. BC and RP analyzed the data and wrote the manuscript. PG and JPV contributed in editing and conceiving the structure of the final manuscript. RP, TM and FNP conceived the study and supervised the research. All authors have read and approved the manuscript.

